# Impaired SARS-CoV-2 mRNA vaccine antibody response in chronic medical conditions: a real-world data analysis

**DOI:** 10.1101/2021.08.03.21261483

**Authors:** Shu-Yi Liao, Anthony N. Gerber, Pearlanne Zelarney, Barry Make, Michael E. Wechsler

**Affiliations:** Department of Medicine, National Jewish Health, Denver, CO; Department of Academic Affairs, National Jewish Health, Denver, CO

## Abstract

This study is to investigate whether certain medical conditions may impair antibody response to the mRNA vaccines. In this unique study, participants were drawn from patients in National Jewish Health, a pulmonary specialty outpatient clinic.Our study highlights fact that 26% of our patients (n=226) who had spike protein ab measured at least 14 days post 2^nd^ vaccine had negative spike protein ab testing. We found interstitial lung disease (ILD) to be an independent risk factor for impaired antibody response. While the exact antibody level that confers protection against SARS-CoV-2 is unknown and there may be other non-B cell-mediated protection (e.g. T cell-mediated), our study raises concerns that SARS-CoV-2 vaccination may not result in protective immunity in all populations and may have implications for some as masking and distancing strategies are abandoned.

## Introduction

While vaccines against COVID-19 have led to dramatic decreases in COVID cases and mortality in the US, the majority of patients in the vaccine trials did not have chronic underlying diseases -patients at the highest risk of morbidity and mortality due to COVID-19 (1). Host factors in these high-risk patients may result in reduced responsiveness to vaccines and “breakthrough” COVID cases. As masking and distancing precautionary measures are lifted, individuals who fail to mount an immune response to the vaccines may change their protective behavior without being aware of their potential vulnerability. NBC News found 125,682 SARS-CoV-2breakthrough infections in the US as of July 30, 2021 (2). A recent report showed that 39 fully vaccinated health care workers had breakthrough infections and neutralizing antibody titers were lower during the peri-infection period than those in matched uninfected controls (3). While a recent study found 46% of transplant patients had no antibody response after two doses of mRNA vaccines (4), no studies to date have investigated the effects of underlying chronic medical conditions on antibody response. This study was to use real-world data to evaluate whether individuals with chronic medical conditions have impaired antibody response to SARS-CoV-2 mRNA vaccines.

## Methods

We used National Jewish Health electronic medical records to identify patients who received 2 doses of SARS-CoV-2 mRNA vaccines between December 16, 2020–July 24, 2021, and had a spike IgG antibody results at least 14 days after the second dose. These tests were clinically ordered by individual physicians at our subspecialty academic medical center focused on chronic respiratory and immune disorders. Anti-SARS-CoV-2 ELISA (EUROIMMUN US, New Jersey) with a recombinant S1 domain of the spike protein was used to measure IgG antibodies to SARS-CoV-2 spike protein. The ratio of the sample optical density (OD) to the OD of a calibration provided with the kit was interpreted as positive (≥0.8) or negative (<0.8) (5). Medical conditions were based on physician diagnosis ICD-10 codes; medications were defined as in a previous study (6). A multivariate logistic regression model was used to identify clinical characteristics associated with the spike IgG protein being negative. Due to the small sample size, overlap propensity score weighting (7) was used to analyze medication effects.

## Results

We identified 226 patients (mean age 62 years; 62% female) who received 2 doses of mRNA vaccines and had antibody testing (Table). BNT162b2 (Pfizer-BioNTech) was administered to 66% of the patients and 34% received mRNA-1273 (Moderna). At a mean (standard deviation) of 62 (35) days, range 14-161 days, after dose 2, 26% had no antibody detected. Negative antibody responses were found in 47 patients receiving the BNT162b2 (Pfizer-BioNTech) vaccination and 11 patients receiving the mRNA-1273 (Moderna) vaccination. Eight patients had documented COVID-19 infection before the vaccination. The percentages of patients without antibodies detected ranged from 14% in chronic obstructive pulmonary disease to 53% in congestive heart failure (CHF) (Table). Multivariate logistic regression (Figure) showed that interstitial lung disease (ILD) (OR 0.21, 95% CI 0.08-0.56), CHF (OR 0.26, 95% CI 0.07-0.98), and use of biologics (anti-IL-5, -IL-6, -IL-12/23, -IL-17, -IgE, -CD20, and -TNF-α inhibitors) /JAK inhibitors (OR 0.17, 95% CI 0.07-0.46) are significant risk factors for negative antibody response adjusted for age, gender, vaccine types, days after vaccination, other medication use and comorbidities. Using overlap propensity score weighting, biologic/JAK inhibitor use is associated with impaired antibody response (OR 0.79, 95% CI 0.68-0.91). We have further separated rituximab (anti-CD20) from the biologics/JAK inhibitors and found that rituximab use is the driver of the above finding (OR 0.57, 95% CI 0.47-0.69) while other biologics/JAK inhibitors use (excluding rituximab) is not associated with negative antibody response. No significant effects on antibody response were observed with other medications including systemic/inhaled corticosteroids, and other immunosuppressants including methotrexate, mycophenolate mofetil, azathioprine, leflunomide, cyclosporine, tacrolimus, mycophenolic acid, sirolimus, and everolimus (data not shown). Given the finding that ILD is an independent risk factor for impaired antibody response, we further investigated the subpopulation with ILD. Among the 87 patients (mean age 65 years; 64% female) evaluated by National Jewish Health ILD specialists, 40 (46%) have negative antibodies. Clinical features such as age, gender, lung function, comorbidity, and medication use were similar in ILD patients with and without antibody responses (Data not shown).

**Table.** Characteristics of the study population (N=226) and antibody response^*^.

| Characteristics | Negative (N=58) | Positive (N=168) |
| --- | --- | --- |
| Age (years), mean (SD) | 65 (14) | 60 (14) |
| Gender |  |  |
| Female | 34 (24%)<sup>†</sup> | 106 (76%) |
| Male | 24 (28%) | 62 (72%) |
| Hypertension | 20 (28%) | 52 (72%) |
| Diabetes | 6 (29%) | 15 (71%) |
| Asthma | 12 (17%) | 58 (83%) |
| COPD | 4 (14%) | 24 (86%) |
| Interstitial lung diseases | 40 (46%) | 47 (54%) |
| Sarcoidosis | 4 (19%) | 17 (81%) |
| Congestive Heart failure | 9 (53%) | 8 (47%) |
| Renal Diseases | 5 (31%) | 11 (69%) |
| Rheumatic diseases | 22 (39%) | 34 (61%) |
| ACEI/ARB | 21 (28%) | 54 (72%) |
| Inhaled corticosteroids | 16 (20%) | 63 (80%) |
| Systemic corticosteroids (exclude tapering dose) | 16 (30%) | 38 (70%) |
| Immunosuppressants | 37 (39%) | 57 (61%) |
| Hydroxychloroquine | 7 (35%) | 13 (65%) |
| Biologics<sup>‡</sup>/JAK inhibitors | 21 (47%) | 24 (53%) |
| Vaccine type |  |  |
| mRNA-1273 (Moderna); | 11 (15%) | 65 (85%) |
| BNT162b2 (Pfizer-BioNTech) | 47 (31%) | 103 (69%) |
| Days after 2<sup>nd</sup> vaccine dose, mean (SD) | 80 (36) | 56 (32) |

\* primary medical conditions and relevant medications
<sup>†</sup> Number and (%) of patients by row
<sup>‡</sup> Biologics: Anti- IL-5, -IL-6, -IL-12/23, -IL-17, -IgE, -CD20, and -TNF- $\alpha$ inhibitors
COPD: chronic obstructive pulmonary disease; ACEI/ARB: angiotensin-converting enzyme inhibitors/angiotensin-receptor blockers
\*Biologics: Anti- IL-5, -IL-6, -IL-12/23, -IL-17, -IgE, -CD20, and -TNF- $\alpha$ inhibitors

**Figure.**
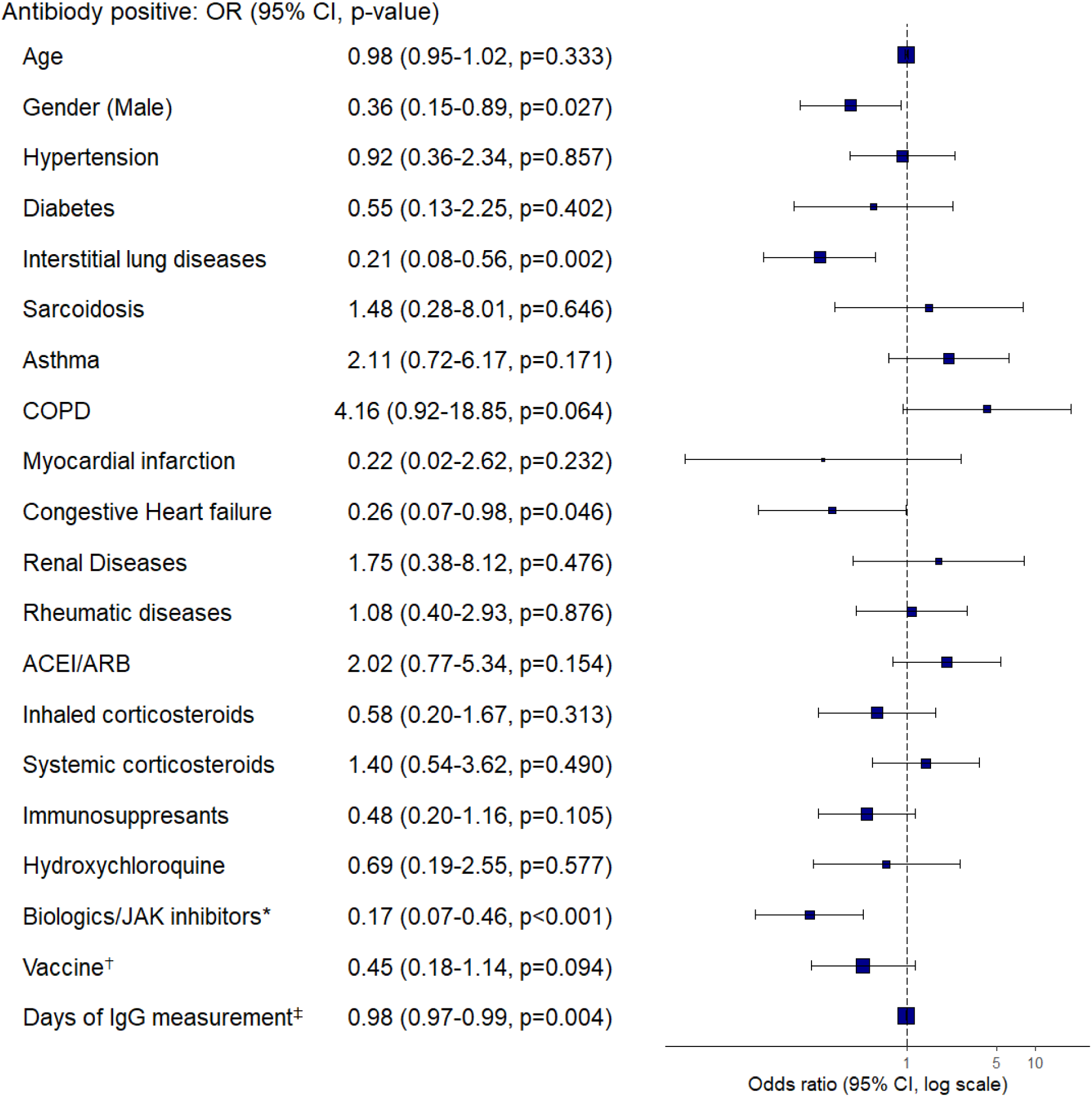
Multivariate logistic regression of the association between antibody response and clinical characteristics. OR: odds ratio; CI: confidence interval; COPD: chronic obstructive pulmonary disease; ACE/ARB: angiotensin-converting enzyme inhibitors/angiotensin-receptor blockers. *Biologics: Anti-IL-5, -IL-6, -IL-12/23, -IL-17, -IgE, -CD20, and -TNF-α inhibitors ^†^BNT162b2 (Pfizer-BioNTech) compared to mRNA-1273 (Moderna) ^‡^Days after 2^nd^ vaccine dose.

## Discussion

It is critical to recognize that vaccination may not necessarily confer immunity, at least as assessed by commercially available spike protein antibody tests, particularly in patients with certain chronic medical conditions and certain medication use. In our patients (79% with chronic pulmonary diseases), 26% had no antibody detected ≥ 14 days after the second dose of mRNA vaccines. While these data show that all vaccinated individuals may be at risk for impaired antibody production, we found that CHF, ILD (independent of their medication use), and biologic/JAK inhibitor use (especially rituximab) are independent risk factors for impaired antibody response. The effect of rituximab use is consistent with a previous study (8). While the exact antibody level that confers protection against SARS-CoV-2 is unknown and there are other markers of B cell and T cell-mediated protection(9), a recent study showed that among fully vaccinated health care workers, the occurrence of breakthrough COVID-19 infection was correlated with the titer of antibodies (3). We have yet to observe the occurrence of COVID-19 infections in these patients with negative antibody responses. However, given the relatively low prevalence of COVID-19 infection in our region, the relatively short time since vaccines were administered, the lack of rigorous follow-up to assess infections, and the recommendations by our physicians to continue mask-wearing and other precautions, this is not surprising. Our study raises concerns that SARS-CoV-2 vaccination may not result in protective immunity in patients with chronic medical conditions. Further studies of immunologic response (including neutralizing antibodies and other markers of B cell and T cell response) and protection in vulnerable populations are needed to define ongoing COVID-19 risk and inform recommendations regarding additional vaccination and behaviors to mitigate risk. Such studies are particularly important with the predicted seasonal and variant-driven spikes in SARS-CoV-2 infection rates.

## Data Availability

N/A

## Acknowledgments

The study was approved by the National Jewish Health Institutional Review Board. The authors would like to thank Joy Zimmer for her help with the electronic health record data query.

## Author Contributions

Dr. Liao had full access to all the data in the study and takes responsibility for the integrity of the data and the accuracy of the data analysis. Concept and design: All authors. Acquisition and interpretation of data: All authors. Drafting of the manuscript: Liao. Critical revision of the manuscript for important intellectual content: All authors. Statistical analysis: Liao. Supervision: Gerber, Make, Wechsler

